# Exploring the association of Obesity on Cold and Warm Autoimmune Hemolytic Anemia in San Joaquin Valley: A Retrospective Cross-Sectional Study

**DOI:** 10.64898/2026.06.11.26355276

**Authors:** Kevin Trong Dao, Rupam Sharma, Rajashree Hariprasad, Remon Indrawes, Wilbur Nelson Montana

**Author notes:** Corresponding author: Kevin Trong Dao, M.D^1^ Kern Medical – UCLA, 1700 Mount Vernon Ave.

## Abstract

The relationship between obesity and specific autoimmune diseases have been well-established, specifically due to obesity’s role in promoting pro-inflammatory states. Although, not much literature had been documented regarding obesity association with AIHA. As such, this study aims to assess any correlations in patients with elevated body mass index (BMI) and autoimmune hemolytic anemia (AIHA). Here we present a retrospective cross-sectional study conducted over a four-year period, across four medical centers during which a new electronic medical record was implemented. The study included 25 patients who have had a previously documented history of AIHA from another facility, DAT positive with indicators of hemolysis, or DAT positive with monomer specific antisera. The patients BMI was recorded at the time of presentation to the hospital. However, for patients with a prior history of AIHA or those transferred from another facility, the BMI that was closest to the time period of when the patient was diagnosed with AIHA was used as an adjunct. Our results show that there is an association of patients with elevated BMI (>25) and AIHA; however, various other confounding variables should be taken into consideration, and further research should be done to establish a causal relationship.

## 1. Introduction

Autoimmune hemolytic anemia (AIHA) is a rare disease caused by autoantibodies that adhere to a patient’s own erythrocytes, thus inducing hemolysis. [1] When a patient’s own bone marrow is unable to keep up with the hemolysis, severe anemia can occur. One unique characteristic of this disease process is that the autoantibodies that cause the hemolytic anemia are active in various thermal temperatures. Immunoglobulin G (IgG) antibodies are more associated with warm AIHA (wAIHA), whereas immunoglobulin M (IgM) or, in some rare instances, anti-PR antigen are associated with cAIHA. [1-3] Numerous prior studies have demonstrated that various risk factors have been known to be associated with warm and cold AIHA. Common risk factors include immunodeficiency, infection, malignancies, autoimmune disorders, pregnancy, etc., all of which can induce immune dysregulation, leading to onset of AIHA. [4-5] Despite warm and cold AIHA sharing similar features of immunoglobulin-mediated destruction, the pathophysiology and laboratory findings differ greatly. One unique factor is that specific types of disease and comorbidities have a higher propensity for patients to develop either cold or warm AIHA and, in some cases, a mixed picture. However, not much is known regarding obesity’s association with AIHA.

As one of the most significant public health challenges in recent years, obesity has affected over hundreds of millions of people worldwide. [6] In fact, since the mid-1970s, the rates of obesity have approximately tripled, with projections noting a future increase of obese patients all throughout the world. [6] The World Health Organization (WHO) itself defines obesity based on the body mass index. [7] Unfortunately, body mass index doesn’t differentiate between lean and fat mass and, as a result, has issues distinguishing body fat distribution as well as the health disease related to fat mass obesity. [6] Despite this, all patients tend to have adipose tissue, which is an energy storage endocrine organ well known to secrete a variety of adipokines, including resistin, adiponectin, leptin, and inflammatory cytokines such as interleukin-6 (IL-6) and tumor necrosis factor-α (TNF-α). [8] Due to this excess, adiposity can promote states of chronic low-grade systemic inflammation. As a result, this can greatly affect the innate and adaptive immune systems, leading to the development of autoimmune disorders. Many studies have depicted strong correlations between obesity and several autoimmune diseases, including but not limited to systemic lupus erythematosus, rheumatoid arthritis, inflammatory bowel disease, Graves’ disease, etc. [9-10] Therefore, we would like to further investigate the effects of obesity on AIHA, specifically whether elevated BMI is commonly observed among patients with AIHA and whether a potential relationship exists between adiposity and AIHA. A discussion regarding the results of the study, limitations, and any and all conclusions will also be held.

## 2. Patient Characteristics and Methods

### 2.1 Study Procedures and Design

This retrospective cross-sectional study was conducted over a four-year period during the implementation of a new electronic medical record (EMR) system. The data was only extracted from four medical centers in the central valley in California. The data was extracted from the EMR using the International Classification of Diseases, 10th Revision (ICD-10) codes. Codes D59.10 (Autoimmune hemolytic anemia, unspecified), D59.11 (Warm autoimmune hemolytic anemia), D59.12 (Cold autoimmune hemolytic anemia), and D59.19 (Other autoimmune hemolytic anemia) were used. Once the patients were identified based on ICD-10 codes, the patients had to meet our inclusion criteria for diagnosis of autoimmune hemolytic anemia (AIHA) (Figure 1). Particularly, the patients either had a documented history of AIHA from another facility or DAT positive with positive indicators of hemolysis or DAT positive with monomer. Regarding the monomers for wAIHA, lab values show DAT positive with IgG with or without C3d, and for cAIHA, DAT is positive for C3d and ± IgM (Figure 1). [4] If the patient had met these criteria, then the patient’s body mass index was recorded at the time he/she presented to the hospital. Although if the patient had a prior history of AIHA that was noted at a previous admission or had a diagnosis of AIHA at another facility, then the BMI closest to the time of diagnosis of AIHA would be used. Lastly the ethical approval to report this study was obtained from the UCLA Kern Medical Center Institutional Review Board.

### 2.2 Patient Characteristics

The median age was 50 years old (age range 24 to 80 years old). The median age of patients with wAIHA is 42 years old, and for cAIHA is 63 years old. Of the 25 patients, 6 are male and 19 were female. The patients had underlying associated medical conditions that are either known to cause autoimmune hemolytic anemia or diseases that severely affect the immune system, such as autoimmune disease, infections, or malignancy. Treatments, particularly medications that suppress the immune system, such as disease-modifying antirheumatic drugs (DMARD), biological drugs, and corticosteroids, were also noted (Table 1).

## 3. Results and Statistical Analysis

Data extraction of the study depicted that out of a total of 25 patients, 19 patients (76%) had warm autoimmune hemolytic anemia (wAIHA), and 6 patients (24%) had cold autoimmune hemolytic anemia (cAIHA). One patient had wAIHA with a BMI <18.5 kg/m^2^. Three patients had a BMI of 18.5-24.9 kg/m^2^, with two patients having wAIHA and one having cAIHA. The common BMI category was the overweight class (BMI 25–29.9 kg/m^2^), with 11 patients noted, comprising 44% of the sample size. Within this category, 8 patients (72.7%) had wAIHA and 3 patients (27.3%) had cAIHA. Ten patients (40%) had a BMI ≥30 kg/m^2^, including 4 patients with BMI 30–34.9 kg/m^2^, three of whom had wAIHA and one had cAIHA. Four patients had a BMI of 35–39.9 kg/m^2^, all of whom had wAIHA. Lastly, two patients (8%) had a BMI >40 kg/m^2^, which was evenly distributed between warm and cold AIHA (Table 2).

Furthermore, among this study, the patient with a normal BMI (18.5–24.9 kg/m^2^) accounted for 12% of the sample size, and within this group, 66.6% had wAIHA and 33.3% had cAIHA. The largest BMI category was overweight (25–29.9 kg/m^2^), representing 44% of all AIHA cases, with 72.7% of said patients having wAIHA and 27.3% having cAIHA. Patients who were noted to have class I obesity (BMI 30–34.9 kg/m^2^) per the WHO [7] comprised 16% of the study, of whom 75% had wAIHA and 25% had cAIHA. Similarly, patients with class II obesity (BMI 35–39.9 kg/m^2^) accounted for 16% of all AIHA cases, with all patients in said category having wAIHA. Lastly, patients with a BMI >40 kg/m^2^ represented 8% of the 25 patients with AIHA and were evenly distributed between wAIHA and cAIHA. Overall, the overweight and obese individuals (BMI ≥ 25 kg/m^2^) constituted the majority of patients with AIHA, accounting for 84% of the study population (Table 2).

Finally, the mean BMI (kg/m^2^) of the patients in our sample size of 25 is noted to be 29.68 ± 2.54 kg/m^2^ with a confidence interval of 95% with the lower bound being 27.14 kg/m^2^ and the upper bound being 32.23 kg/m^2^. For a confidence interval of 99%, the lower bound is 26.34 kg/m^2^, whereas the upper bound is noted to be 33.02 kg/m^2^ with a standard deviation of 6.48 kg/m^2^ from the mean with a variance of 42.03 (Table 3). For wAIHA, 19 patients were noted with a mean BMI of 29.27 ± 2.64 kg/m^2^ with 95% confidence, whereas for 99% confidence, the mean BMI is 29.27 ± 3.475 kg/m^2^ with a standard deviation of 5.88 kg/m^2^. As for cAIHA, 6 patients were noted with a mean BMI of 30.99 ±6.36 kg/m^2^ with 95% confidence, whereas for 99% confidence, the mean BMI is 30.99 ± 8.37 kg/m^2^ with a standard deviation of 7.96 kg/m^2^.

Overall, 21 of 25 patients (84%) were overweight or obese (BMI ≥25 kg/m^2^), suggesting a high prevalence of elevated BMI among patients with AIHA. Warm AIHA predominated across nearly all BMI categories, accounting for 76% (19/25) of the study.

## 4. Discussion

As both an immunological and hematological disease, cold and warm AIHA are both fairly rare hematologic disorder resulting in the immune system mistakenly destroying red blood cells. [1-3] Due to the fact that this disease is an autoimmune disorder, many factors that trigger the immune system can result in this disease. However, one factor that isn’t well documented is body mass index (BMI) associations with AIHA. As a widely used measurement to estimate body mass, BMI values can have various clinical implications. Particularly, obesity itself is associated with increased adiposity, which can promote chronic states of low-grade systemic inflammation, which can increase chances of autoimmune disease. [9-10]

Further investigation of other associated medical conditions indicates that there is a likelihood that elevated BMI may contribute to conditions such as autoimmune disease, malignancy and infection, which themselves are associated with AIHA. The proposed concept would be that BMI would predispose patients to having autoimmune disease, infections, malignancies, etc., which would increase the chances of the patients having AIHA. For example, one prospective cohort study of 238,130 women reported evidence that obesity was associated with an 85% increased risk of developing SLE (HR 1.85, 95% CI 1.17–2.91). [11] This is a crucial factor as various meta-analysis study had noted that approximately 11.8% of SLE patients had developed AIHA. [12] The relationship between obesity and malignancies is another example of development of AIHA. In fact, one study had noted that patients who had a BMI greater than 25 had higher rates of malignancy and had noted that a reduction of 10% of weight loss or greater can possibly reduce cancer risk. [13-15] Furthermore, the same study had noted at least 13 cancers linked to overweight and obese patients. More specifically, patients had higher chances of developing diffuse large B-cell lymphoma (DLBCL) and hematologic malignancies. [13-15]

This key association of obesity and diffuse large B-cell lymphoma (DLBCL) is key since said cancer has been noted to be a cause of cAIHA. [16] Lastly, there is strong evidence to support obesity’s association with an increased risk of infections with a clear dose response. One large multicohort study of over 540,000 participants had noted that patients with class 1 obesity were at approximately 1.5 times higher risk, class 2 obesity was at approximately 2 times higher risk, and class 3 obesity was at approximately 3 times higher risk. [17] These infections would, as a result, further predispose patients to developing either warm or cold AIHA. Overall, these indirect causes of how overweight and obese patients could develop other underlying conditions that predispose patients to developing either warm or cold AIHA.

With regards to how BMI can directly cause AIHA, it should be noted that BMI has had some associations with autoimmune disease prior, with a recent meta-analysis and systematic review noting an approximately 41% higher risk of incidence of autoimmune disease in obese individuals. [18-19] Although there are instances where there is an inverse association seen with other autoimmune diseases such as systemic sclerosis and Sjögren’s syndrome. [18] There are various proposed mechanisms as to how this can occur. Beyond the chronic low-grade inflammatory state caused by adipose-mediated immune response, other studies have reported leptin’s ability to enhance activation of both the adaptive and innate immune systems. [20-21] Another proposed possibility that another study had noted would be the blocking of a key regulator of Treg cells (FOXP3), resulting in inhibition of their proliferation, thereby promoting a proinflammatory response. [22]

In our study, the average BMI of all AIHA patients was 29.68 kg/m^2^, and for wAIHA patients, the mean was 29.27 kg/m^2^, whereas for cAIHA was 30.99 kg/m^2^ (Table 3). This depicts a pattern of patients that are noted to be overweight in all AIHA and wAIHA groups, with the cAIHA group meeting the criteria for class 1 obesity per the WHO. [7] This could be due to a variety of proposed mechanisms that could result in the patient developing AIHA; however, further studies should be done to ensure a causal relationship.

Lastly, it should also be noted that there are other factors that could have played a role in the patients having AIHA. Within our sample size, 6 patients were male, whereas 19 patients were female, reflecting the prevalence of autoimmune diseases within specific genders in the greater population. Gender has been highly associated with autoimmune disease due to the X chromosome, sex hormone production, and other regular genes that are either more predominant or lacking in one gender versus the other. [24-26] Age has been noted to play a factor in autoimmunity as well [26-27], with our population having a median age of 50 years old; specifically, patients with wAIHA had a median age of 42 years old, whereas patients with cAIHA had a median age of 63 years old (Table 1). It should be highlighted that the median was reported rather than the mean since a majority of AIHA studies report medians due to the age distributions, which are skewed, particularly in wAIHA, which has a bimodal distribution with peaks in younger adults with autoimmune conditions. [4] As such, it is very possible that these factors also played a part in patients developing AIHA, depicting a more multi variable process.

## 5. Study Limitations

In this study there are several limitations that should be considered. Most importantly, there are various confounding variables, such as infections, malignancies, and underlying autoimmune diseases, that should be noted since each variable in and of itself can be independently associated with autoimmune hemolytic anemia (AIHA). Additionally, various biases should be noted, particularly selection bias, since the study population was derived from only four medical centers, which predominantly serve underserved patient populations and may not be representative of the broader AIHA population. Measurement bias should also be noted since BMI values were obtained from different facilities using varying equipment. Furthermore, the BMI was recorded closest to the time of AIHA diagnosis if the patient was diagnosed with AIHA from another facility rather than a measurement obtained at the time of diagnosis itself. Also, the value ofBMI itself has inherent limitations as a measure of adiposity, as it does not distinguish between fat mass and lean body mass. Lastly, since AIHA is a fairly rare disease in and of itself, the sample size of our study is relatively small, which limits the statistical power and may reduce the ability to detect meaningful associations between BMI and AIHA subtypes.

## 6. Conclusion

While causality cannot be established in this retrospective cross-sectional study, overweight and obesity were common among patients with AIHA, with 84% of patients having a BMI ≥25 kg/m^2^. These findings support further investigation into the relationship between adiposity, immune dysregulation, and AIHA in larger prospective studies.

## Data Availability

All data produced in the present work are contained in the manuscript

## Ethical Approval

Ethical approval to report this study was obtained from the Kern Medical Institutional Review Board; approval ID: 24069

## Acknowledgments

None.

