## Supplementary material for "Exploring the association of Obesity on Cold and Warm Autoimmune Hemolytic Anemia in San Joaquin Valley: A Retrospective Cross-Sectional Study": Figure 1.

Figure 1. Flow-chart depicting the step wise fashion of the study’s' inclusion criteria

Patients were screened from our facility database (n = 4) for 4 years

Patients that were screened

Screening was based on ICD-10 code

International Classification of Diseases, 10th Revision (ICD-10) Included

D59.10 = Autoimmune hemolytic anemia, unspecified

D59.11 = Warm autoimmune hemolytic anemia

D59.12 = Cold autoimmune hemolytic anemia

D59.19 = Other autoimmune hemolytic anemia

Inclusion Criteria

The patients were included based on a documented history of AIHA from another facility

or

DAT positive with positive indicators of hemolysis such as elevated indirect bilirubin, lactate dehydrogenase, decreased haptoglobin, etc.

or

DAT positive with monomer specific antisera (IgG, C3d, IgM) [11]

Total Patients Included (n = 25)
