## Supplementary material for "Exploring the association of Obesity on Cold and Warm Autoimmune Hemolytic Anemia in San Joaquin Valley: A Retrospective Cross-Sectional Study": Table 1.

Table 1. Key Clinical and Laboratory Characteristics

| Age Range /Sex | Auto Immune Hemolytic Anemia (AIHA) | Body Mass Index (BMI) | Lowest Hemoglobin during admission (Grams/Deciliter) | Highest Hemoglobin after treatment during admission (Grams/Deciliter) | Associated Medical conditions* | Treatment** | Outcome |
| --- | --- | --- | --- | --- | --- | --- | --- |
| 25-30 Male | Warm | 17.6 | 4.6 g/dL | 8.1 g/dL | Human Immunodeficiency Virus | Blood Transfusion | The patient was discharged with resolution and hemoglobin stabilized |
| 55-60 Female | Cold | 21.34 | 4.5 g/dL | 9.9 g/dL | Systemic Lupus Erythematosus | Blood Transfusion, Rituximab, Prednisone, IV Iron Infusions | The patient had improvement with treatment and still continuing management |
| 25-30 Female | Warm | 35.98 | 5.5 g/dL | 10.5 g/dL | Antiphospholipid Syndrome and Systemic Lupus Erythematosus | IV Immunoglobulin, Prednisone, Blood Transfusion | The patient was discharged with resolution and hemoglobin stabilized |
| 75-80 Male | Cold | 29.3 | 5.2 g/dL | 8.2 g/dL | Mycoplasma Pneumonia | Blood Transfusion | The patient was discharged with resolution and hemoglobin stabilized |
| 45-50 Female | Warm | 36 | 5.7 g/dL | 11.4 g/dL | Latent Tuberculosis | Rituximab, Methylprednisolone, Prednisone, Blood Transfusion | The patient was discharged with resolution and hemoglobin stabilized |
| 70-75 Female | Cold | 34.7 | 6.3 g/dL | 9.3 g/dL | Septic Shock | Blood Transfusion | The patient unfortunately passed away |
| 50-55 Female | Warm | 25.6 | 4.2 g/dL | 14.9 g/dL | Antiphospholipid Syndrome and Systemic Lupus Erythematosus | Prednisone, Blood Transfusion | The patient left against medical advice and didn’t follow up |
| 40-45 Female | Warm | 23.48 | 6.5 g/dL | 8.4 g/dL | Syphilis, Hepatitis C, Systemic Lupus Erythematosus, Rheumatoid Arthritis | Blood Transfusion | The patient was discharged with resolution and hemoglobin stabilized |
| 20-25 Female | Warm | 33.86 | 6.9 g/dL | 9.0 g/dL | Immune Thrombocytopenia, Sars Cov 2 | Prednisone | The patient was discharged with resolution and hemoglobin stabilized |
| 50-55 Female | Warm | 26.4 | 4.7 g/dL | 9.2 g/dL | Systemic Lupus Erythematosus, Antiphospholipid Syndrome, Bacteremia/Sepsis | Blood Transfusion | The patient was discharged with resolution and hemoglobin stabilized |
| 20-25 Female | Warm | 30.05 | 4.6 g/dL | 10.7 g/dL | Systemic Lupus Erythematosus | Blood Transfusion, IV Immunoglobulin, Prednisone, Folic Acid, Rituximab | The patient was discharged with resolution and hemoglobin stabilized |
| 60-65 Male | Cold | 28.6 | 3.6 g/dL | 8.4 g/dL | Chronic Lymphocytic Leukemia and Septic Shock | Blood Transfusion | The patient was transfered to higher level of care via air ambulance |
| 55-60 Female | Warm | 36.98 | 9.5 g/dL | 11.9 g/dL | Helicobacter Pylori, Systemic Sclerosis (Scleroderma) | Rutixmab, Prednisone | The patient was discharged with resolution and hemoglobin stabilized |
| 35-40 Female | Warm | 35.74 | 11 g/dL | n/a | Systemic Lupus Erythematosus | Prednisone | The patient was discharged with resolution of underlying admission |
| 35-40 Female | Warm | 26.45 | 10 g/dL | 11.5 g/dL | None | IV Immunoglobulin, Prednisone, Rituximab, Mycophenalate | The patient was discharged with resolution and hemoglobin stabilized |
| 45-50 Male | Warm | 25.45 | 12.9 g/dL | 14.2 g/dL | Sars Cov 2 | Prednisone | The patient was discharged with resolution and hemoglobin stabilized |
| 50-55 Female | Warm | 40.94 | 10.5 g/dL | 12.5 g/dL | Mixed Connective Tissue Disease, Systemic Lupus Erythematosus, Hashimotos Thyroiditis | Blood Transfusion for treatment years prior, Metotrexate, Folic Acid | The patient was discharged with resolution and hemoglobin stabilized |
| 65-70 Female | Warm | 26.94 | 5.9 g/dL | 8.9 g/dL | Immune Thrombocytopenia (ITP), Helicobacter pylori | Prednisone, Rituximab, Blood Transfusion | The patient was discharged with resolution and hemoglobin stabilized |
| 60-65 Female | Warm | 30.04 | 5.6 g/dL | 9.6 g/dL | None | Methylprednisolone, Prednisone, Blood Transfusion | The patient was discharged with resolution and hemoglobin stabilized |
| 40-45 Female | Warm | 28.87 | 6.2 g/dL | 10.2 g/dL | Undifferentiated Connective Tissue Disease, Systemic Lupus Erythematosus, Streptococcus pneumoniae | IV Immunoglobulin, Prednisone, Folic acid, Hydroxychloroquine, Blood Transfusion | The patient was discharged with resolution and hemoglobin stabilized |
| 25-30 Male | Warm | 25.01 | 10.8 g/dL | 12.7 g/dL | Human Immunodeficiency Virus (HIV)/Acquired Immunodeficiency Syndrome (AID), Syphillis | Prednisone | The patient was discharged with resolution and hemoglobin stabilized |
| 30- 35 Female | Cold | 46.37 | 8.8 g/dL | n/a | None | Supportive Measures | The patient was discharged with resolution and hemoglobin stabilized |
| 35-40 Female | Warm | 29.89 | 6.8 g/dL | 8.6 g/dL | Epstein–Barr virus and Mycoplasma Pneumoniae | Blood Transfusion, Prednisone, Folic Acid | The patient was transferred to higher level of care and was loss to follow up |
| 45- 50 Male | Warm | 20.97 | 7.1 g/dL | 10.4 g/dL | Ulcerative Colitis, Multifocal Pneumonia, Urosepsis, Monoclonal Gammopathy of Undetermined Significance, Mycobacterium Avium | Prednisone | The patient was discharged with resolution and hemoglobin stabilized |
| 60-65 Female | Cold | 25.67 | 6.6 g/dL | 9.4 g/dL | Celiac Disease, Plasma Cell Dyscrasia or Lymphoplasmacytic Lymphoma (Loss to Followup for Diagnosis) | Rituximab, Blood Transfusion | The patient was discharged with resolution and hemoglobin stabilized |

*Associated medical conditions were autoimmune disease, infections, and/or malignancies due to such conditions having a prevalence to cause auto immune hemolytic anemia (AIHA)

** Treatment include treatment for auto immune hemolytic anemia (AIHA) but also included immunosuppressive medications used to treat other autoimmune disease noted in associated medical conditions since they can also affect AIHA

*** n/a = Not available because the patient was not being treated for AIHA and a repeat complete blood count was not needed or the patient had refused a repeat complete blood count
