## Supplementary material for "Exploring the association of Obesity on Cold and Warm Autoimmune Hemolytic Anemia in San Joaquin Valley: A Retrospective Cross-Sectional Study": Table 2.

Table 2. Body Mass Index (BMI) corresponding with Autoimmune Hemolytic Anemia

| Body Mass Index (BMI) | Warm Autoimmune Hemolytic Anemia (n = 19) (wAIHA) | Cold Autoimmune Hemolytic Anemia (n = 6) | Total Autoimmune Hemolytic Anemia (tAIHA) with corresponding BMI | % of patients with wAIHA with corresponding BMI (a*/c***) | % of patients with cAIHA with corresponding BMI (b**/c***) | % of patients with tAIHA with corresponding BMI compared to total patients (c***/d****) |
| --- | --- | --- | --- | --- | --- | --- |
| <18.5 kg/m² | 1 | 0 | 1 | 100 % | 0 % | 4 % |
| 18.5-24.9 kg/m² | 2 | 1 | 3 | 66.6 % | 33.3 % | 12 % |
| 25-29.9 kg/m² | 8 | 3 | 11 | 72.7 % | 27.3 % | 44 % |
| 30-34.9 kg/m² | 3 | 1 | 4 | 75 % | 25 % | 16 % |
| 35-39.9 kg/m² | 4 | 0 | 4 | 100 % | 0 % | 16 % |
| >40 kg/m² | 1 | 1 | 2 | 50 % | 50 % | 8 % |

*a = Corresponds to the number of patients with Warm Autoimmune Hemolytic Anemia (wAIHA) with the corresponding BMI as noted in the column

**b = Corresponds to the number of patients with Cold Autoimmune Hemolytic Anemia (cAIHA) with the corresponding BMI as noted in the column

***c = Corresponds to the number of patients with Total Autoimmune Hemolytic Anemia (Both Warm and Cold)(tAIHA) with the corresponding BMI as noted in the column

****d = Corresponds to the total number of patients with Total Autoimmune Hemolytic Anemia (Both Warm and Cold)(tAIHA)(n=25)
