## Supplementary material for "Exploring the association of Obesity on Cold and Warm Autoimmune Hemolytic Anemia in San Joaquin Valley: A Retrospective Cross-Sectional Study": Table 3.

Table 3. Statistical Analysis of Body Mass Index and AIHA

| Total AIHA Patient Count (n) | 25 |
| --- | --- |
| Mean BMI (μ) of all AIHA patients with Confidence Interval of (95 %) | 29.68 ±2.54 kg/m² = (27.14, 32.23) |
| Mean BMI (μ) of all AIHA patients with Confidence Interval of (99 %) | 29.68 ±3.34 kg/m² = (26.34, 33.02) |
| Standard Deviation from Mean BMI of all AIHA patients (σ) | 6.48 kg/m² |
| Variance of all AIHA patients (σ2) | 42.03 |
| Warm AIHA Patient Count (n) | 19 |
| Mean BMI (μ) of Warm AIHA patients with Confidence Interval of (95 %) | 29.27 ±2.64 kg/m² = (26.62, 31.91) |
| Mean BMI (μ) of Warm AIHA patients with Confidence Interval of (99 %) | 29.27 ±3.47 kg/m² = (25.79, 32.74) |
| Standard Deviation from Mean BMI of Warm AIHA patients (σ) | 5.88 kg/m² |
| Variance of Warm AIHA patients (σ2) | 34.58 |
| Cold AIHA Patient Count (n) | 6 |
| Mean BMI (μ) of Cold AIHA patients with Confidence Interval of (95 %) | 30.99 ±6.36 kg/m² = (24.62, 37.35) |
| Mean BMI (μ) of Cold AIHA patients with Confidence Interval of (99 %) | 30.99 ±8.37 kg/m² = (22.61, 39.36) |
| Standard Deviation from Mean BMI of Cold AIHA patients (σ) | 7.96 kg/m² |
| Variance of Cold AIHA patients (σ2) | 63.38 |
